# Clinical Validity of the diagnostic criteria for metabolic-associated fatty liver disease: a real-world experience

**DOI:** 10.1101/2020.08.20.20176214

**Authors:** Yasser Fouad, Zienab M Saad, Hend M Moeness, Ehab M Abdel-Raheem, Yasser Abdelghani, Nasr M Osman, Wafaa Abdelhameed, Alaa M Mostafa, Dina Attia

## Abstract

**Background and Aims:** To eliminate the anti-definition of non-alcoholic fatty liver disease (NAFLD), positive clinical criteria for metabolic associated fatty liver disease are recently proposed. In this study, we examine the validation and utilization of these criteria.

**METHODS:** Two cohorts of 316 consecutive patients were recruited, including 242 patients previously diagnosed to have NAFLD and 74 patients with concomitant NAFLD and chronic hepatitis C (CHC) The validity of the proposed criteria for MAFLD, namely presence of hepatic steatosis with one of three criteria, overweight/obesity, diabetes or evidence of metabolic dysregulation was assessed. Fibrosis was assessed using, fibrosis-4 (FIB-4) and NAFLD fibrosis score (NFS). The impact of MAFLD on the clinical outcomes in CHC patients was also investigated.

**Results:** The clinical criteria captured 240 patients (99.2%). 215 (88.8%) met either overweight or diabetes and 25 (10.3%) met the presence of 2 criteria of metabolic dysfunction. In patients, with dual etiologies, in the multivariable analysis adjusting for age, sex, BMI, ALT, AST and diabetes, the presence of MAFLD were significantly associated with increase high FIB-4 score of fibrosis (Odds ratio [95% confidence interval], 3.77 [1.49–9.48], P < 0.005) when compared to those with MAFLD only.

**CONCLUSION:** The proposed criteria for diagnosis of MAFLD is well validated and easily applicable to the entire spectrum of disease including non-obese subjects. Patients with lean MAFLD have favorable metabolic and fibrosis characteristics compared to their obese counterpart, while patients with concomitant MAFLD and CHC had severe metabolic and fibrosis characteristics compared to patients with MAFLD alone.

## Introduction

Metabolic associated fatty liver disease (MAFLD) (formerly known as non-alcoholic fatty liver disease) is the most common liver disease that affects about 25–30% of the global population(1–3). The MAFLD is a multisystem disease with wide consequences, as it not only increases the risk of liver cirrhosis and failure and hepatocellular carcinoma (HCC), but also increase the risk for various extra-hepatic complications such as cardiovascular disease, chronic kidney disease, type 2 diabetes, osteoporosis, and some types of cancers(4).

The pathogenesis of the disease is complex and involves a dynamic interaction between environmental factors and genetic basis(5, 6), with evidence of shared genetics between MAFLD and other metabolic diseases(7). Although, classically, linked to obesity, there is an emerging evidence that a considerable proportion of patients are non-obese, as called lean MAFLD (8). However, the characteristics of this group of patients are not well characterised, particularly in Caucasian white population.

Worryingly, the Middle East region has the highest prevalence of MAFLD globally, in parallel with dramatic nutrition transition and high rates of physical inactivity in this region(9, 10). However, the current “anti-definition” and linking to alcohol, the disease with labeling “NAFLD” represents a major lacuna that hinders the clinical and research progress(11).

Adding to the complexity, the Middle East region, and particularly Egypt had one of the highest prevalence of hepatitis C worldwide(12, 13). The current definition of NAFLD that is based on exclusion of other diseases, was very problematic and limits the ability to assess the actual magnitude of both problems and proper management of liver diseases that patients have.

Recently, an international consensus proposed new diagnostic criteria for MAFLD that could potentially help to eliminate the current “anti-definition”(2). The criteria are based on evidence of hepatic steatosis, in addition to one of the following three criteria, namely overweight/obesity, presence of type 2 diabetes mellitus, or evidence of metabolic dysregulation. However, the validity and robustness of these proposed criteria in a real-world cohorts are still unknown.

Thus, in this work, we examined the validity of the proposed criteria for MAFLD in a real-life cohort, characterize the characteristics of lean MAFLD and finally determine the characteristics of patients with MAFLD/HCV dual etiologies. Here, we report that the proposed criteria for diagnosis of MAFLD is well validated and easily applicable and capture the entire spectrum of disease including non-obese subjects.

## Methods

### Study cohort

This is a prospective two centre cross-sectional study included 242 patients previously diagnosed with non-alcoholic fatty liver disease who met the inclusion criteria were consecutively included in the study. Patients were recruited from the outpatient clinics of Minia University Hospitals and Beni-Suef University Hospitals, Egypt. Individuals with alternative diagnoses were excluded including those with excess alcohol intake (>10 g per day for women; and >20 g per day for men), chronic viral hepatitis (hepatitis B and hepatitis C), decompensated liver disease, known active malignancy or an alternative cause for fatty liver. The validity of the novel criteria proposed by the international expert panel to diagnose MAFLD was investigated (2). The novel criteria for diagnosis of MAFLD are shown in Figure 1. g per day for women; and > 20 g per day for men),

**Figure 1:**
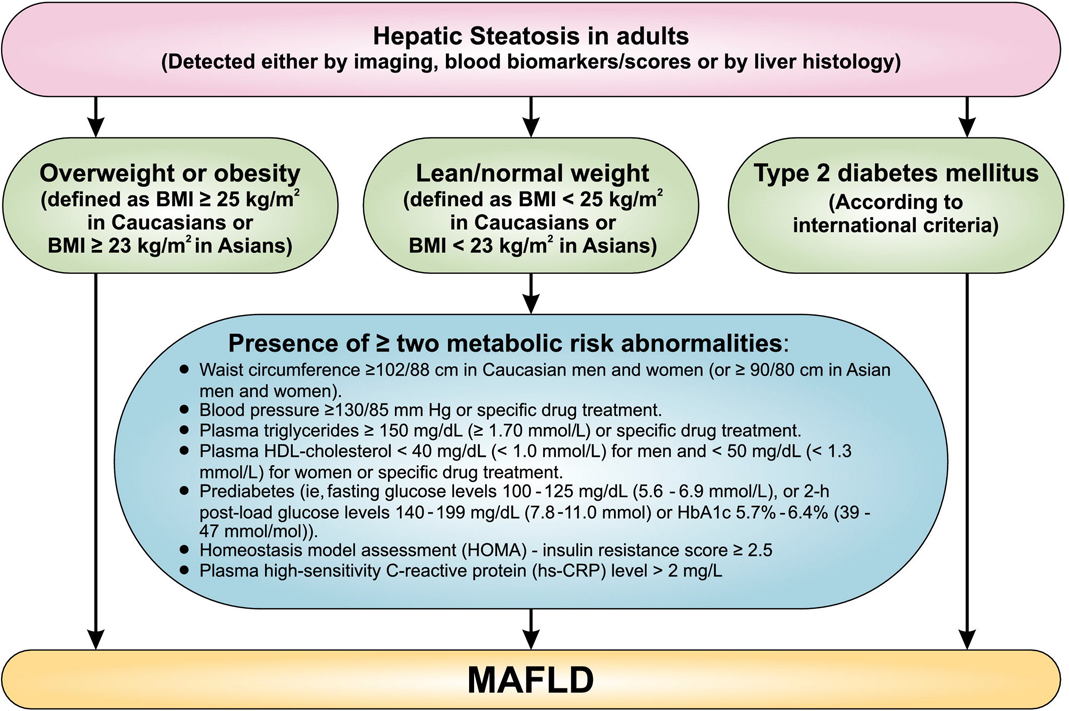
The novel proposed diagnostic criteria for metabolic associated fatty liver disease (Adopted from Eslam M, et al 2020) (2)

To investigate the impact of dual eitology on liver fibrosis, a second cohort of patients with dual pathology HCV and MAFLD criteria was included (n = 78), based on the criteria mentioned above, apart from allowing for including HCV-positive patients.

The study was conducted in accordance to the ethics of good clinical practice and Helsinki Declaration after approval of the local ethical committee. A written informed consent was obtained from all patients.

### Clinical and laboratory assessments

All participants underwent a clinical history using a standardized questionnaire that collected information on age, gender, smoking, and alcohol intake. Weight (in kilograms) and height (in centimetres) were measured and the body mass index (BMI) was calculated and expressed as kg/m^2^. Waist circumference was taken at the midpoint between the lowest rib and the top of the iliac crest and measured in centimetres. Hypertension was defined as a resting blood pressure of ≥130/80 mm Hg, or having any antihypertensive medication prescribed. Type 2 diabetes mellitus (T2DM) was defined as a fasting plasma glucose value ≥ 126 mg/dL or having any antidiabetic medication prescribed. The homeostasis model assessment (HOMA-IR) was calculated as (fasting serum insulin (μU m^−1^) × fasting serum glucose (mmol ^1−^))/22.5. Fasting blood samples were taken in the morning for all participants and analyses for full blood count, liver biochemistry, lipid profile, fasting glucose and iron studies were performed.

In the dual eitology cohort, HCV was diagnosed using HCV RNA levels, carried out using the COBAS AmpliPrep /COBAS TaqMan HCV Quantitative Test, version 2.0, with a lower limit of quantification of 20 IU/m.

### Ultrasound

All patients received an abdominal ultrasonography using (Acuson X300, Siemens, USA) and CH5–2 probe. A single experienced examiner in each centre screened all patients in each centre and diagnosis of fatty liver was based upon ultrasound diagnostic criteria of fatty liver (14–16).

### Statistical analysis

For descriptive statistics, values were expressed as mean ± standard deviation, median and interquartile range as appropriate. The Mann-Whitney non-parametric test was used to obtain significance. For categorical variables, data were presented as frequency (percentage) and p-values. Comparisons of distributions between groups was assessed using Fisher’s exact test. Multivariable regression modelling with backward elimination was undertaken to test independent associations with significant fibrosis and FIB-4 score in the dual aetiologies (MAFLD and HCV). All statistical analyses were undertaken in Statistical analyses were performed using the statistical software package SPSS for Windows, version 21(SPSS, Chicago, IL).

## Results

### The proposed criteria for diagnosis of MAFLD is well validated patients and applicable

We explored the validity of new “positive” criteria for diagnosis of MAFLD in a cohort of 242 patients with previously diagnosed NAFLD. Noticeably, in this cohort, the diagnostic criteria were captured in 240 patients (99.2%). The overweight criterion was met in 212 (87.6) patients, and either overweight or diabetes were met in 215 (88.8%) patients. The presence of 2 criterion of metabolic dysfunction were met in 25 (10.3%) patients. As, only two subjects (0.8%) who do not meet the MAFLD criteria, we were not able to investigate for their characteristics. In total, the newly proposed criteria are valid and capture virtually patients previously diagnosed to have NAFLD.

### Lean MAFLD patients have favourable metabolic and fibrosis characteristics

The prevalence and characteristics of subjects with lean MAFLD are still not completely known, particularly in other ethnicities apart from Caucasian. Thus, we next thought to investigate this in our cohort. Interestingly, 100 (41.3%) patients were non-obese (defined as BMI <30 kg/m^2^), while 30 (12.3%) patients were lean (defined as BMI <25 kg/m^2^),

The baseline characteristics of the nonobese MAFLD patients compared to their obese counterpart are depicted in **Table 1**. Non-obese MAFLD patients had favourable metabolic profile, as by definition they had significantly lower BMI and waist circumference, as well as lower fasting blood sugar and HOMA-IR, while lipid profile and other characteristics were not significantly compared to their obese counterparts.

**Table 1:** baseline characteristics of MAFLD patients

|  | Non-obese<br>n = 100 | Obese<br>n = 142 | P-value |
| --- | --- | --- | --- |
| **Clinical parameters** |  |  |  |
| Age (years) (mean ± SD) | 48 ± 12 | 48 ± 11 | 0.75 |
| Female/Male N (%) | 68/32 (68/32) | 105 /37 (74/26) | 0.31 |
| Diabetes mellitus N (%) | 20 (20%) | 37 (26.1%) | 0.29 |
| Hypertension (%) | 18 (18%) | 38 (26.8%) | 0.12 |
| BMI (kg/m<sup>2</sup>) | 26 ± 3 | 35 ± 4.2 | <0.001 |
| Waist circumference (cm) | 102.1 | 102.45 | <0.001 |
| **Laboratory parameters** |  |  |  |
| Hemoglobin (g/dL) | 13.0 ± 1.8 | 13 ± 1.7 | 0.9 |
| Total leucocytic count (/mm<sup>3</sup>) | 7.0 ± 2.4 | 7.0 ± 2.2 | 0.73 |
| Platelet count (10<sup>9</sup>/L) | 259 ± 76 | 278 ± 83 | 0.04 |
| INR | 1.08 ± 0.1 | 1.07 ± 0.9 | 0.84 |
| ALT (U/L) | 30 ± 19 | 28 ± 20 | 0.31 |
| AST (U/L) | 34 ± 21 | 30 ± 25 | 0.04 |
| Bilirubin (total) mg/dL | 0.2 ± 0.1 | 0.2 ± 0.2 | 0.2 |
| Creatinine (mg/dL) | 0.9 ± 0.3 | 0.9 ± 0.3 | 0.84 |
| Fasting blood sugar (mg/dL) | 73 ± 18 | 97 ± 23 | 0.08 |
| Random blood sugar(mg/dL) | 131 ± 69 | 162 ± 134 | 0.02 |
| Hemoglobin A1c (%) | 9 ± 1.4 | 7.2 ± 1.4 | 0.71 |
| HOMA-IR score | 4.8 (3.3-8.5)<br>8.64±9.48 | 6.6 (4.1-14.9)<br>11.56±12.05 | 0.01 |
| Total cholesterol (mg/dL) | 205 ± 62<br>180 (159-238) | 187 ± 60<br>180 (143-216) | 0.15 |
| Triglycerides (mg/L) | 92 ± 59 | 145 ± 72 | 0.13 |
| HDL-cholesterol (mg/L) | 44 ± 11 | 44 ± 10 | 0.92 |
| LDL-cholesterol (mg/dL) | 127 ± 72 | 118 ± 65 | 0.21 |
| **Non-invasive scores for fibrosis** |  |  |  |
| NFS | 0.457 ± 1.522 | 0.950 ± 1.507 | 0.01 |
| FIB-4 | 1.370 ± 1.026 | 1.145 ± 0.828 | 0.14 |

BMI, body mass index; INR, international normalization ratio; ALT, alanine aminotransferease AST, aspartate aminotransferase; hemoglobin A1C, glycated hemoglobin; HOMA-IR score, homeostatic model assessment of insulin resistance; HDL, high density lipoproteins; LDL, low density lipoproteins; NFS, non-alcoholic fatty liver disease score fibrosis score; FIB-4, fibrosis-4 index, N, number; SD, standard deviation; L, liter; U, unit; Kg, kilogram; cm, centimeters; g/dL, gram/ deciliter; mg, milligram.

Non-obese MAFLD patients also have lower fibrosis, as measured by NFS (p=0.010), while FIB-4 was not significantly different (**Table 1**), while this association was less profound when we applied the low and high cut-off for each score. (**Table 2**)

**Table 2:** Baseline characteristics of MAFLD patients with single versus dual pathology

|  | MAFLD<br>n = 242 | MAFLD+HCV<br>n = 67 | P-value |
| --- | --- | --- | --- |
| **Clinical parameters** |  |  |  |
| Age (years) (mean ± SD) | 48 ± 12 | 49 ± 13 | 0.37 |
| Female / Male N (%) | 172 /70 (71/29) | 42 /33(57/43) | 0.62 |
| Diabetes mellitus N (%) | 57 (23.6%) | 19 (28.4%) | 0.43 |
| Hypertension (%) | 56 (23.1%) | 16 (23.9%) | 0.87 |
| BMI (kg/m<sup>2</sup>) | 31 ± 5.6 | 30 ± 4.3 | 0.26 |
| BMI> (30 kg/m<sup>2</sup>) | 142 (58.7%) | 32 (47.8%) | 0.13 |
| Waist circumference (cm) | 102.2 ± 13.7 | 103.3 ± 11.4 | 0.17 |
| **Laboratory parameters** |  |  |  |
| Hemoglobin (g/dL) | 13 ± 1.7 | 13 ± 1.7 | 0.49 |
| Total leucocytic count (/mm<sup>3</sup>) | 7.0 ± 2.3 | 8.1 ± 13.4 | 0.07 |
| Platelet count (10<sup>9</sup>/L) | 270 ± 80 | 248 ± 66 | 0.06 |
| ALT (U/L) | 29 ± 19 | 32 ± 16 | 0.01 |
| AST (U/L) | 32 ± 24 | 35 ± 19 | 0.09 |
| Creatinine (mg/dL) | 0.9 ± 0.3 | 0.9 ± 0.2 | 0.7 |
| Random blood sugar(mg/dL) | 149 ± 113 | 128 ± 54 | 0.36 |
| Hemoglobin A1c (%) | 7.3 ± 1.4 | 7.6 ± 1.7 | 0.5 |
| HOMA-IR score | 10 ± 11 | 18 ± 13 | 0 |
| Total cholesterol (mg/dL) | 194 ± 61 | 224 ± 58 | <0.001 |
| Triglycerides (mg/dL) | 148 ± 67 | 153 ± 50 | 0.22 |
| HDL-cholesterol (mg/dL) | 44 ± 11 | 45 ± 11 | 0.4 |
| LDL-cholesterol (mg/dL) | 122 ± 68 | 149 ± 73 | 0 |
| **Non-invasive scores for fibrosis** |  |  |  |
| NFS | 0.743 ± 1.530 | 0.843 ± 1.811 | 0.05 |
| FIB-4 | 1.240 ± 0.921 | 1.476 ± 1.187 | 0.12 |

BMI, body mass index; INR, international normalization ratio; ALT, alanine aminotransferease AST, aspartate aminotransferase; hemoglobin A1C, glycated hemoglobin; HOMA-IR score, homeostatic model assessment of insulin resistance; HDL, high density lipoproteins; LDL, low density lipoproteins; APRI, Aspartate aminotransferase to platelets ratio index; NFS, non-alcoholic fatty liver disease score fibrosis score; FIB-4, fibrosis-4 index, N, number; SD, standard deviation; L, liter; U, unit; Kg, kilogram; m<sup>2</sup>, meter square; cm, centimeters; g/dL, gram/ deciliter.

### Dual aetiologies; MAFLD and HCV

Having confirmed the robustness of the MAFLD diagnostic criteria, we next explored the impact of HCV/MAFLD dual aetiologies compared to MAFLD alone. Baseline characteristics of the dual aetiologies are described in **Table 3**. Interestingly the HOMA-IR, total cholesterol and LDL-cholesterol were found to be are significantly higher in patients with dual aetiologies compared to MAFLD alone (p< 0.001, for all comparisons). Notably, patients with the dual aetiologies have also significantly higher fibrosis compared to those with MAFLD alone, assessed by FIB-4 score (p = 0.001) (**Table 4**). This association remained significant after adjusting for gender, AST and ALT in multiple regression analysis (Odds ratio [95% confidence interval], 3.77 [1.49–9.48], P < 0.005) (**Table 5**).

**Table 3:** association between the different fibrosis scores and the MAFLD

|  | **Non-obese**<br>**N= 100** | **Obese**<br>**N=142** | **P-value** |
| --- | --- | --- | --- |
| **NFS** |  |  |  |
| <-1.455 | 9 (9%) | 8 (5.6%) | 0.45 |
| >-1.455 | 91 (91%) | 134 (94.4%) |  |
| **FIB-4** |  |  |  |
| <1.3 | 70 (70%) | 103 (72.5%) | 0.67 |
| >1.3 | 30 (30%) | 39 (27.5%) |  |

APRI, Aspartate aminotransferase to platelets ratio index; NFS, non-alcoholic fatty liver disease score fibrosis score; FIB-4, fibrosis-4 index.

**Table 4:** association between the different fibrosis scores and the dual etiologies

|  | **MAFLD** | **MAFLD + HCV** | **P-value** |
| --- | --- | --- | --- |
| **NFS** |  |  |  |
| <-1.455 | 17 (6.9%) | 9 (12.2%) | 0.15 |
| >-1.455 | 228 (93.1%) | 65 (87.8%) |  |
| **FIB-4** |  |  |  |
| <1.3 | 175 (71.4%) | 37 (50%) | 0 |
| >1.3 | 70 (28.6%) | 37 (50%) |  |

MAFLD, metabolic associated fatty liver disease, HCV, hepatitis C virus; APRI, Aspartate aminotransferase to platelets ratio index; NFS, non-alcoholic fatty liver disease score fibrosis score; FIB-4, fibrosis-4 index.

**Table 5:** Multiple regression analysis for dual etiologies associated with severe fibrosis assessed by FIB-4

|  | P-value | Odds ratio | 95% CI |  |
| --- | --- | --- | --- | --- |
|  |  |  | Upper | lower |
| Dual etiologies | 0 | 3.33 | 1.61 | 6.87 |
| Gender | 0.02 | 0.43 | 0.22 | 0.88 |
| BMI | 0.34 | 1.03 | 0.97 | 1.09 |
| DM | 0.07 | 0.53 | 0.26 | 1.06 |
| ALT | <0.001 | 0.94 | 0.92 | 0.97 |
| AST | <0.001 | 1.16 | 1.12 | 1.2 |

HCV Ab, hepatitis C virus antibodies; BMI, body mass index; INR, international normalization ratio; ALT, alanine aminotransferease AST, aspartate aminotransferase; DM, diabetes mellitus.

In total, patients with dual HCV/MAFLD aetiologies present with severe metabolic and fibrosis characteristics.

## Discussion

This is the first evaluation of the diagnostic accuracy of the novel criteria proposed by the international expert panel to diagnose MAFLD (2) in a real-world cohort of patients previously diagnosed with NAFLD. We demonstrate that the criteria were easy to apply and fit to the diagnosis in nearly all patients (99%), have an excellent diagnostic property with virtually all patients (99%) were identified, with sensitivity (98.4%) and negative predictive value (96.1%). The fact that there is no need to depend on exclusion of other causes of chronic liver disease enhances the feasibility of these criteria.

The new name, MAFLD, was distinguished by several benefits, including accurate and easy diagnosis of the disease in addition to providing appropriate health care and avoidance of stigmatization, trivialization and proper catalyst to increase funding and heath plan action (11). It was not long before the new nomenclature, MAFLD, was published until the researchers grabbed it. Applying the MAFLD criteria in studying a cohort of patients with COVID-19 infection in China was published recently (17). The investigators found that the presence of MAFLD increased the risk obesity on severity of illness with COVID-19infection. In the same context, other researcher, in a multicenter study, found a severe form of Covid-19 in younger patients with MAFLD in the preliminary analysis (18). This finding was attributed to the systemic and hepatic immune response caused by MAFLD contributing to cytokine storm in patients with COVID-19.

Another intriguing finding in our study, non-obese MAFLD patients represent more than 40% of all patients with a higher ratio than other areas in the world and 12.3% were lean MAFLD. Patients with lean MAFLD have favourable metabolic characteristics and fibrosis scores compared to their obese counterparts. The newly proposed criteria for MAFLD incorporating a consideration of metabolic health would be of highl utility in better characterisation of this group of patients, in a manner extend beyond the measurement of obesity by BMI. From clinical standpoint, the proper identification of non-obese MAFLD is crucial. First, While, a recent study suggested that non obese may even benefit from weight reduction as the obese patients with NAFLD (19), the non-obese especially lean individuals may not be easily convinced with the concept of losing weight and exercise which are of pivotal role in treatment of MAFLD. Secondly non obese individuals are less likely to get much attention from their doctors regarding their fatty liver diseases or associated metabolic dysfunction. Applying the recent proposed MAFLD criteria easily identified these patients with accuracy.

Though the pathogenesis of lean MAFLD is not completely understood, a recent study that is shaped by differential metabolic adaptation, that is mediated by increased bile acids and Farnesoid X receptor (FXR) activity compared to the obese counterparts(20). Lean MAFLD patients also tends to have distinct microbiota profile and frequency of genetic variants, such as Transmembrane 6 superfamily 2 and (TM6SF2) interferon lambda 3 (IFNL3) compared to obese subjects(20–22).

Noticeably, the new MAFLD criteria allowed us to better define patients with dual aetiologies. Herein, we characterised the nature of (MAFLD-HCV) instead of exclusion of HCV in previous NAFLD name leading to a drop of large number of patients with actual metabolic dysfunction especially in countries with high prevalence of HCV like Egypt. When comparing MAFLD patients with patients with dual pathology (MAFLD-HCV) we found not only a significant increase in the metabolic parameters but also a severer fibrosis in patients with dual pathology. This association remained significant after adjusting gender, ALT and AST in multiple regression analysis.

Identification of MAFLD and dual pathologies may lead to a notable change in the liver disease map globally. In a recent report revising data of 2.14 million liver related death, NAFLD was diagnosed in 9% of patients while HCV was diagnosed in 26% and alcoholic liver disease was disease including non-obese subjects and to identify a previously largely overlooked group who are having dual etiologies of MAFLD with other liver diseases.

Identification of MAFLD and dual pathologies may lead to a notable change in the liver disease map globally. In a recent report revising data of 2.14 million liver related death, NAFLD was diagnosed in 9& of patients while HCV was diagnosed in 26& and alcoholic liver disease was diagnosed in 25% who died of chronic liver diseases (23). We think that health care and research could get benefit from distinguishing metabolic associated fatty liver diseases and dual pathologies for better management and decreasing the mortality.

The study could have some limitation, the cohort was recruited in a tertiary centre, so it might be subject of selection bias and lack of liver biopsy hinders the details assessment of histological features. Further studies would be required to ascertain that these proposed criteria would achieve the claimed high utility in other communities with different genetic backgrounds, environmental factors, and health care systems than those in the Egypt.

In conclusion, the diagnostic criteria proposed by the international consensus panel for the diagnosis of MAFLD EDCT are easy to apply, and have excellent diagnostic properties, which make them useful for routine clinical practice, large population-based epidemiological studies or for, clinical trials. In addition, application of these criteria allowed to capture the entire spectrum of disease including non-obese subjects and to identify a previously largely overlooked group who are having dual etiologies of MAFLD with other liver diseases.

## Data Availability

the data will be available

## Notes

**Conflict of interest:** The authors have nothing to disclose.

### Competing Interest Statement

The authors have declared no competing interest.

### Funding Statement

No funding

### Author Declarations

Approaval by faculty of Medicine, Minia University Approaval number: 328-11/2019

## References

1. Eslam M, Sanyal AJ, George J, International Consensus P. MAFLD: A Consensus-Driven Proposed Nomenclature for Metabolic Associated Fatty Liver Disease. Gastroenterology 2020.May;158(7):1999–2014.e1. doi:10.1053/j.gastro.2019.11.312

2. Eslam M, Newsome PN, Anstee QM, Targher G, Gomez MR, Zelber-Sagi S, Wong VW, et al. A new definition for metabolic associated fatty liver disease: an international expert consensus statement. 2020 Apr 8:S0168–8278(20)30201–4. doi:10.1016/j.jhep.2020.03.039

3. Younossi Z, Anstee QM, Marietti M, Hardy T, Henry L, Eslam M, George J, et al. Global burden of NAFLD and NASH: trends, predictions, risk factors and prevention. Nature reviews Gastroenterology & hepatology 2018;15:11.

4. Byrne CD, Targher G. NAFLD: a multisystem disease. Journal of hepatology 2015;62:S47-S64.

5. Eslam M, Valenti L, Romeo S. Genetics and epigenetics of NAFLD and NASH: clinical impact. Journal of hepatology 2018;68:268–279.

6. Bayoumi A, Grønbæk H, George J, Eslam M. The Epigenetic Drug Discovery Landscape for Metabolic-associated Fatty Liver Disease. Trends in Genetics 2020. Available online 28 March 2020 https://doi.org/10.1016/j.tig.2020.03.003

7. Eslam M, George J. Genetic contributions to NAFLD: leveraging shared genetics to uncover systems biology. Nature Reviews Gastroenterology & Hepatology 2020 Jan;17(1):40–52. doi:10.1038/s41575-019-0212-0.

8. Chen F, Esmaili S, Rogers G, Bugianesi E, Petta S, Marchesini G, Bayoumi A, et al. Lean NAFLD: a distinct entity shaped by differential metabolic adaptation. Hepatology 2020 Apr;71(4):1213–1227. doi:10.1002/hep.30908.

9. Guthold R, Stevens GA, Riley LM, Bull FC. Worldwide trends in insufficient physical activity from 2001 to 2016: a pooled analysis of 358 population-based surveys with 1.9 million participants. Lancet Glob Health 2018;6:e1077-e1086.

10. Azizi F, Hadaegh F, Hosseinpanah F, Mirmiran P, Amouzegar A, Abdi H, Asghari G, et al. Metabolic health in the Middle East and north Africa. The Lancet Diabetes & Endocrinology 2019 Nov;7(11):866–879. doi:10.1016/S2213-8587(19)30179-2.

11. Fouad Y, Waked I, Bollipo S, Gomaa A, Ajlouni A, Attia D. What's in a name? Renaming “NAFLD” to “MAFLD”. Liver International 2020 Apr 17. doi:10.1111/liv.14478. Online ahead of print.

12. Kandeel A, Genedy M, El..Refai S, Funk AL, Fontanet A, Talaat M. The prevalence of hepatitis C virus infection in Egypt 2015: implications for future policy on prevention and treatment. Liver International 2017;37:45–53.

13. Khattab MA, Ferenci P, Hadziyannis SJ, Colombo M, Manns MP, Almasio PL, Esteban R, et al. Management of hepatitis C virus genotype 4: recommendations of an international expert panel. Journal of hepatology 2011;54:1250–1262.

14. Kim SH, Lee JM, Kim JH, Kim KG, Han JK, Lee KH, Park SH, et al. Appropriateness of a donor liver with respect to macrosteatosis: application of artificial neural networks to US images--initial experience. Radiology 2005;234:793–803.

15. Dasarathy S, Dasarathy J, Khiyami A, Joseph R, Lopez R, McCullough AJ. Validity of real time ultrasound in the diagnosis of hepatic steatosis: a prospective study. J Hepatol 2009;51:1061–1067.

16. Hernaez R, Lazo M, Bonekamp S, Kamel I, Brancati FL, Guallar E, Clark JM. Diagnostic accuracy and reliability of ultrasonography for the detection of fatty liver: a meta-analysis. Hepatology 2011;54:1082–1090.

17. Zheng KI, Gao F, Wang XB, et al. Obesity as a risk factor for greater severity of COVID-19 in patients with metabolic associated fatty liver disease. Metabolism. 2020;154244. doi:10.1016/j.metabol.2020.154244

18. Zhou YJ, Zheng KI, Wang XB, Yan HD, Sun QF, Pan KH, Wang TY, Ma HL, Chen YP, George J, Zheng MH. Younger patients with MAFLD are at increased risk of severe COVID-19 illness: A multicenter preliminary analysis, Journal of Hepatology (2020), doi: https://doi.org/10.1016/j.jhep.2020.04.027.)

19. Wong VW, Wong GL, Chan RS et al. Beneficial effects of lifestyle intervention in non patients with non alcoholic fatty liver disease. J Hepatol 2018; 69: 1349–56)

20. Chen F, Esmaili S, Rogers G, Bugianesi E, Petta S, Marchesini G, Bayoumi A, et al. Lean NAFLD: A Distinct Entity Shaped by Differential Metabolic Adaptation. Hepatology. 2020;71(4):1213–1227. doi:10.1002/hep.30908

21. Eslam M, Hashem AM, Leung R, Romero-Gomez M, Berg T, Dore GJ, Chan HL, et al. Interferon-λ rs12979860 genotype and liver fibrosis in viral and non-viral chronic liver disease. Nature communications 2015;6:1–10.

22. Petta S, Valenti L, Tuttolomondo A, Dongiovanni P, Pipitone RM, Cammà C, Cabibi D, et al. Interferon lambda 4 rs368234815 TT> δG variant is associated with liver damage in patients with nonalcoholic fatty liver disease. Hepatology 2017;66:1885–1893.

23. Paik JM, Golabi P, Younossi Y, Mishra A, Younossi ZM. Changes in the Global Burden of Chronic Liver Diseases From 2012 to 2017: The Growing Impact of Nonalcoholic Fatty Liver Disease [published online ahead of print, 2020 Feb 11]. Hepatology. 2020;10.1002/hep.31173.

